# Trajectories of Perceived Negative Impact in Parents of Individuals with Autism Spectrum Disorders from 9 to 25 years of age

**DOI:** 10.1101/2022.02.23.22271423

**Authors:** Kourtney Christopher, Rebecca Elias, Catherine Lord

## Abstract

Caregivers raising a child with autism experience increased parental burden, though many have positive experiences as well. Perceived negative impact, a form of parental burden, is the degree to which a caregiver reports negative financial, social, and emotional experiences associated with having a child with a disability. This longitudinal study defined parental perceived negative impact trajectory classes across time, determined predictors, and explored the relationship between functional adult outcomes and impact class. Participants (*n* = 209) were comprised of caregivers whose child received an ASD diagnosis or had developmental delay. Latent class growth modeling defined three trajectory classes (e.g., low [*n* = 68, 32.54%], medium [*n* = 98, 46.89%], and high [*n* = 43, 20.57%]). Regressions revealed the low impact trajectory class had more caregivers who were racial minorities, less educated, and more socially supported. Membership in the high negative impact class was associated with increased childhood hyperactivity, irritability, autism symptomology, and poor adaptive skills. The low negative impact trajectory class was associated with young adults achieving more functional outcomes. Overall negative impact declined over time all for all classes, though never completely subsided. Possible implications for clinical practices are discussed

---

Although there are many positive experiences associated with raising a child on the autism spectrum (Kayfitz et al., 2010; King et al., 2016), the extant literature largely has focused on the greater burden that caregivers of children with autism spectrum disorder (ASD) experience, relative to neurotypical children (Bonis, 2016; Estes et al., 2013; McStay et al., 2019; Picardi et al., 2018). Caregiver burden is often conceptualized in two ways. Objective burden refers to difficulties within family relationships, constraints in finances, social, leisure, and work activities, including daily tasks (Picardi et al., 2018). Subjective burden refers to the loss of hopes, dreams, and expectations, increased mental health conditions, and embarrassment in social contexts (Angold et al., 1998; Picardi et al., 2018). Among parents who have a child with ASD, research has primarily focused singularly on subjective burden, commonly called parental stress (Barroso et al., 2018; Hayes & Watson, 2013; Tsermentseli & Evangelia-Chrysanthi, 2019; Zaidman-Zait et al., 2014, 2017). We argue that a holistic conceptualization of caregiver burden is warranted through the measurement of subjective and objective burden together. Several studies, including two describing the present sample, have examined these two facets of autistic caregiver burden through measures of perceived negative impact (Angold et al., 1998; Bishop et al., 2007; Carr & Lord, 2012; Marsack-Topolewski & Church, 2019; Picardi et al., 2018).

## Measurement of Parental Perceived Negative Impact

Parental perceived negative impact assesses the degree to which raising a child with a disability affects parent experiences, including financial, social, emotional, and physical burdens (Messer et al., 1996). The Child and Adolescent Impact Assessment (CAIA; Messer et al., 1996) is a psychometrically sound, semi-structured interview administered to parents to measure perceived negative impact. The CAIA, included in studies of child psychiatric conditions (e.g., Great Smoky Mountain Study [Costello et al., 1996]), indicated that increased levels of child psychopathology and related impairment were linked to high levels of parental perceived negative impact (Angold et al., 1996). Recent studies sought to extend this research by measuring perceived negative impact in parents of children with ASD. Although it is not a focus of this study, increased parental burden and hence stress, has downstream effects on child development for both children with and without ASD (Crum & Moreland, 2017; Jones et al., 2021; Raina et al., 2004; Shine & Perry, 2010).

## Perceived Negative Impact in ASD

To our knowledge, only two studies have examined predictors of parental perceived negative impact in ASD. Both studies used the Early Diagnosis Longitudinal Sample (Bishop et al., 2007; Carr & Lord, 2012), which we also describe in this paper. Bishop and colleagues (2007) explored perceived negative impact among caregivers whose children were nine years old. Parent-reported restricted and repetitive behaviors (RRBs) and low adaptive skills in school-aged children were predictive of increased parental perceived negative impact (Bishop et al., 2007). Further, lower levels of parental perceived negative impact were reported by less educated minority mothers, mothers with more children, and those who felt socially supported compared to more educated mothers with and without minority status, mothers with fewer children, and mothers with less social support (Bishop et al., 2007). Carr and Lord (2012) extended these findings by examining changes longitudinally between childhood and early adolescence. Parental perceived negative impact increased with time, similar to the caregiver stress literature, which describes increased parental stress around the teenage years (Pastor-Cerezuela et al., 2015; Rodriguez et al., 2019). Results also replicated the previous finding that racial minority mothers reported less perceived negative impact relative to White mothers (Bishop et al., 2007; Carr & Lord, 2012).

These findings of less burden reported by mothers of racial minorities stand in stark contrast to the parental stress literature noting increased stress among racial minority parents (Williams et al., 2019). For example, African-American parents of children with autism reported more stress and were more likely to engage in varied coping strategies than Euro-American parents (Williams et al., 2019). Moreover, African-American families with low family resilience levels were more likely to report higher stress levels than White and Hispanic families (Kim, Dababnah, & Lee, 2020). Both of these studies only investigated stress (i.e., subjective burden) in families of autistic children during mid-childhood. Thus, there remain a number of questions in assessing racial minority caregivers’ perceptions of perceived negative impact (i.e., objective and subjective) from childhood to emerging adulthood. Longitudinal analyses provide an opportunity to examine race-related differences in how parents of autistic youth experience burden over time.

## Implications of Trajectories of Negative Impact and Adult Outcomes

Parents of children with autism report higher levels of stress and perceived negative impact that persists throughout children’s early years (Bishop et al., 2007; Rodriguez et al., 2019; Picardi et al., 2018; Zaidman-Zait et al., 2017), and increases into adolescence (Anderson, 2008; Carr & Lord, 2012; Gray 2002; Rodriguez et al., 2019). Recent research has shown that most young adults with autism, at least in one British and one U.S. sample, remain heavily tied to and, in most cases, dependent on their families (McCauley et al., 2020; Stringer et al., 2020). Thus, we need more information about parents’ and caregivers’ perceptions of the implications of having an adult child with autism. Trajectory modeling provides a unique opportunity to examine predictors of parental perceived negative impact from childhood and its relationship to functional outcomes in emerging adulthood.

While it is difficult to define *good* adult outcomes (Howlin et al., 2013; Lounds Taylor, 2017), positive adult outcomes should be described within reasonable expectations given someone’s abilities (Howlin et al., 2013; Pickles et al., 2020; Taylor et al., 2015). McCauley and colleagues (2020) created functional outcome groupings in this longitudinal sample by quantifying the number of functional outcomes related to three categories: autonomy, friends, and purpose. The number of functional outcomes attained was related to participant characteristics such as race, emotional and behavioral problems, autism severity, daily living skills, and happiness. One aim of the present study is to explore the association between parental perceived negative impact and functional outcomes for autistic adults.

The current study examined a longitudinal cohort of participants with autism and developmental delay from childhood through emerging adulthood. The aims were to: 1) identify and describe distinct trajectory classes associated with perceived negative impact for caregivers across time, 2) examine caregiver and participant characteristics that predict membership in the trajectory classes, and lastly, 3) determine if trajectory class was related to the number of adult functional outcomes met by participants.

## Methods

### Participants

Participants were caregivers and their children with an ASD diagnosis or developmental delay (see Lord et al. 2020). Individuals were assessed at three points in time between the ages of nine and 25. The original sample from North Carolina and Illinois (*n* = 213) consisted of consecutive referrals for possible autism for children aged 2. The sample increased by 40 (total *n* = 253) with the addition of a cohort of similar referrals for autism in later childhood from Michigan, with longitudinal data used in this study from early adolescence through adulthood. Inclusion criteria required completion of a caregiver interview about perceived negative impact (i.e., the CAIA) during at least one time point, resulting in a final sample of 209 participants. Attrition was statistically significant in racial minority families with less than a four-year college degree (*p* < .001) but not significantly associated with the site, gender, ASD diagnosis, or Intelligence Quotient (IQ) of participants at age nine.

Consistent with previous research using this sample (Bishop et al., 2007; Carr & Lord, 2012), age bins associated with assessment time points were defined as follows: Time 1: childhood (*n*= 158; *M*_*age*_ = 10.00 years, *SD* = 1.13, *range =* 6.83-11.92), Time 2: early adolescence (*n*= 120; *M*_*age*_ = 13.57 years, *SD* = 1.54, *range =* 12.00-17.92), and Time 3: emerging adulthood (*n*= 116; *M*_*age*_ = 19.61 years, *SD* = 1.31, *range =* 18.00-24.03). Across time participants were primarily White (75-83%), male (81-8%), and had an autism diagnosis (79-84%).. Other more recent diagnoses for those who never received an ASD diagnosis were Intellectual Disability, Attention Deficit Hyperactivity Disorder, or various Learning Disabilities (see McCauley et al., 2020). Most often, mothers (82-91%) reported on perceived negative impact; 76% of the caregivers were married at the first study encounter (see Table 1 for further demographic information).

**Table 1.** Demographic information for the sample by time

|  | Time 1<br>*n* =158 | Time 2<br>*n* =120 | Time 3<br>*n* =116 |
| --- | --- | --- | --- |
| *Family Characteristics* |  |  |  |
| Interview Informant (%) |  |  |  |
| Mother | 82.28 | 91.45 | 83.81 |
| Other Caregiver | 17.72 | 8.55 | 16.19 |
| Education (%) |  |  |  |
| At least four-year college degree | 51.90 | 55.83 | 62.07 |
| Race (%) |  |  |  |
| White | 74.68 | 77.50 | 82.76 |
| Racial minorities | 25.32 | 22.50 | 17.24 |
| Site (%) |  |  |  |
| North Carolina | 54.43 | 58.33 | 43.10 |
| Chicago | 41.77 | 23.33 | 37.93 |
| Michigan | 3.80 | 18.33 | 18.97 |
| Maternal age<br>(*M*, *SD*) | 39.49 (5.61) | 43.26 (6.30) | 49.85 (6.06) |
| Marital Status (%) |  |  |  |
| Married | 75.82 | 66.67 | 60.22 |
| CAIA |  |  |  |
| Total Negative Impact<br>(*M*, *SD*) | 21.56 (10.28) | 23.29 (9.79) | 17.62 (9.94) |
| Feelings of support (%) | 68.59 | 64.66 | 72.07 |
| *Child characteristics* |  |  |  |
| Age (*M*, *SD*) | 10.00 (1.13) | 13.57 (1.54) | 19.61 (1.31) |
| Male (%) | 83.54 | 80.83 | 81.90 |
| ASD (%) | 83.54 | 74.17 | 79.31 |
| NVIQ Above 70 (%) | 41.77 | 47.50 | 44.83 |

*Note.* <9% data was missing from Times 1-3

### Procedure

As part of the longitudinal study, participants and caregivers completed assessments comprised of interviews, questionnaires, and direct testing. Data was collected at two face-to-face visits (i.e., childhood and emerging adulthood) and a telephone interview (i.e., early adolescence). Clinicians with experience in ASD conducted autism-specific assessments, including research-reliable diagnostic measures. Best estimate clinical diagnoses were made on the basis of all information (see McCauley et al., 2020). Informed consent was collected before each visit, and the Institutional Review Board approved the research.

### Measures

#### Parental Perceived Negative Impact

The CAIA (Messer et al., 1996) is a 32-item semi-structured interview assessing both objective and subjective aspects of caregiver burden associated with the child’s condition (i.e., autism or developmental delay). The interview began with a summary of the child’s symptoms and then focused on the impact of those symptoms. “Burden” scores ranged from 0-3, with three being the greatest negative impact for most items. A total of 23 negative impact items were analyzed, ranging from 0-48 (with a score of 48 being the most negatively affected) across four domains. Three of the four domains assessed objective burden with inquiries about the impact on a caregiver’s social life, leisure activities, and financial burden due to having a child with a disability. The fourth domain assessed subjective impact by inquiring about the caregiver’s mental health. Following Bishop et al. (2007), a CAIA question about social support was dichotomized (e.g., yes/no support) because of a skewed distribution.

#### Autism Symptomatology Measures

The Autism Diagnostic Observation Schedule-Second Edition (ADOS-2; Lord et al., 2012) and the Autism Diagnostic Interview-Revised (ADI-R; Lord et al., 1994) were administered by research-reliable clinicians blinded to previous diagnoses. The ADOS-2 has domain Calibrated Severity Scores (CSS) ranging from 1-10 for the total score, as well as Social Affect (ADOS-SA) and RRB domain (ADOS-RRB). The ADI-R is a caregiver interview used to evaluate autism in social, communication, and RRB domains. Increased scores within each domain in each measure indicate higher levels of autistic symptoms. For the purpose of this study we focused on the ADI-RRB domain.

#### Cognitive and Adaptive Skills Measures

Cognitive ability (including verbal [VIQ] and non-verbal IQ [NVIQ]) was assessed using a hierarchy of instruments based on the participant’s cognitive level, expressive language level, and age (see McCauley et al., 2020). Mullen Scale for Early Learning (MSEL; Mullen, 1995), Wechsler Abbreviated Scale of Intelligence (WASI; Wechsler, 1999), Wechsler Intelligence Scale for Children Third Edition (WISC-IV; Wechsler, 2003), or Differential Ability Scales-Second Edition (DAS-2; Elliott, 1990). Adaptive skills were assessed using the Vineland Adaptive Behavior Scales-Second Edition (VABS-II; Sparrow et al., 2005). The VABS-II is a standardized parent interview of adaptive functioning, which yields domain scores in Communication, Daily Living Skills, and Socialization. These domain scores provide an adaptive behavior composite standard score. For this study, we examined the Daily Living (VABS DL) domain in childhood (usually about age nine).

#### Maladaptive Behaviors

The Aberrant Behavior Checklist (ABC; Aman et al., 1985) is a 58-item parent-report questionnaire frequently used to assess behavior difficulties in children and adults with developmental disabilities. Caregivers rated their child’s maladaptive behaviors in the past four weeks on a four-point scale ranging from 0-3, with higher scores indicating greater severity. The ABC contains five subscales: Irritability, Social Withdrawal, Stereotypy, Hyperactivity, and Inappropriate Speech. This study focused on the Irritability (ABC-I) and Hyperactivity (ABC-H) subscales, as Stereotypy, Social Withdrawal, and Inappropriate Speech overlap with the core features of autism. A mean was generated for participants who completed multiple ABC questionnaires near a target time point (i.e., two questionnaires in childhood).

#### Functional Outcomes in Autism

Global functional outcomes were measured using a count variable ranging from 0-3 based on the number of individual functional outcomes met in the areas of autonomy, social relationships, and purpose (McCauley et al., 2020). For more cognitively able adults, positive outcomes were defined as living independently, having employment, and having at least one true friend. For less cognitively able adults, positive outcomes were defined as having at least one friend they enjoy seeing regularly, daily living skills at or above an 8-year-old level, and regular activities outside the house such as work or volunteering (McCauley et al., 2020).

### Data Analytic Plan

#### Aim 1: Identifying Distinct Trajectory Classes of Parental Perceived Negative Impact

To determine trajectory classes of parental perceived negative impact, latent class growth modeling was conducted in STATA 16 using the traj plugin (Jones & Nagin, 2007; 2013; Jones et al., 2001). Models were estimated using full-information maximum likelihood. Participants with incomplete data were included in the analysis under the missing-at-random assumption. Model selection was an iterative process that estimated the number of trajectory classes and each trajectory class’s shape. The lowest Bayesian Information Criterion (BIC) was used to indicate a good model fit.

#### Aim 2: Comparing Trajectory Classes and Predicting Class Membership from Childhood and Parental Characteristics

Data was analyzed using IBM SPSS Statistics Version 27. Multinomial logistic regression was conducted using the low impact trajectory class as a reference group to determine which participant and caregiver characteristics were significant predictors of perceived negative impact class. Selected predictors at age 10 included ADOS-SA, ADOS-RRB, ADI-RRB, ABC-I, ABC-H, VABS-DL, VIQ, presence of autism, race, caregiver education, social support, and marital status. Based on past findings (Bishop et al., 2007; Carr & Lord, 2012), increased behavioral problems (i.e., high irritability and hyperactivity), higher RRBs, lower adaptive skills, and low caregiver social support ratings, but not cognitive ability were hypothesized to predict membership in a trajectory class associated with high perceived negative impact (Bishop et al., 2007; Carr & Lord, 2012).

#### Aim 3: Parental Perceived Negative Impact Trajectory Class and Number of Outcomes

Ordinal logistic regression was performed to investigate whether the perceived negative impact trajectory class was related to the number of functional outcomes adults met. The number of functional outcomes was the dependent variable, and trajectory class was the independent variable. Given the parental stress literature that notes fewer positive outcomes for autistic adults who are living at home with families under chronic stress (DesChamps et al., 2020), the expectation was that those in the highest perceived negative impact class would meet fewer functional outcomes as adults.

## Results

### Trajectory Classes

Latent class growth modeling yielded three parental perceived negative impact classes. (BIC = -1445.26). Figure 1 visually displays the trajectory classes based on the probability that each observation falls into one of the following latent classes: 1) a low impact class that decreased in negative impact over time (*n* = 68, 32.54% of the sample, linear shape), 2) a medium impact class that fluctuated in negative impact over time (*n* = 98, 46.89% of the sample, quadratic shape), and lastly, 3) a high impact class that gradually decreased in negative impact over time (*n* = 43, 20.57%, linear shape). Average posterior probabilities for participants belonging to distinct classes resulted in 0.84 for the low impact class, 0.79 for the medium impact class, and 0.82 for the high impact class, with >0.70 indicating a good fit (Nagin, 2005). Similar to data reported for the full sample in Carr and Lord, the medium impact class increased in negative impact during adolescence but then decreased into early adulthood. In contrast, the other classes comprising about 50% of the sample had negative impact scores that steadily declined across time.

**Figure 1.**
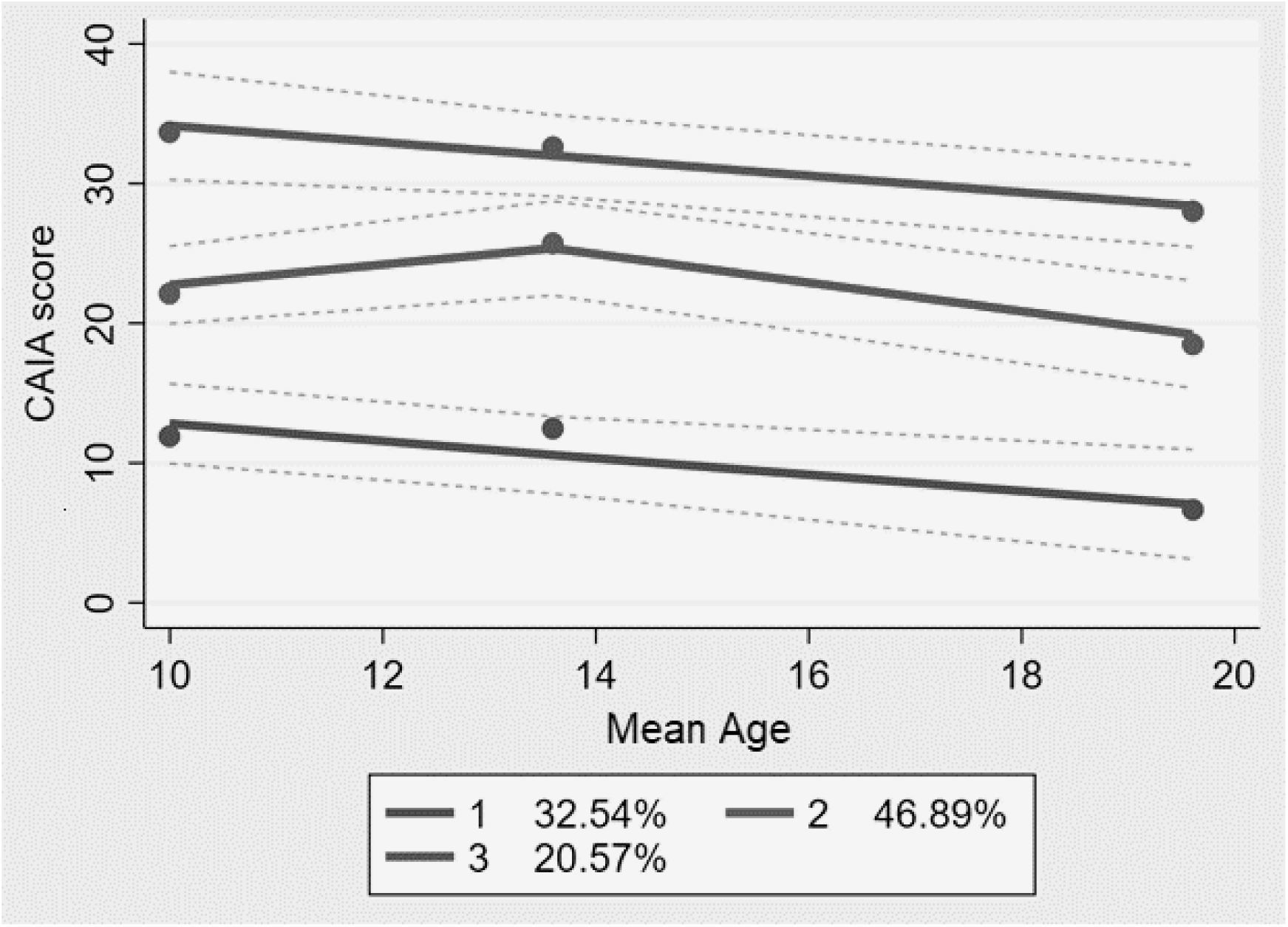
Trajectory Classes of Perceived Negative Impact

### Predicting Class Membership from Child and Parental Characteristics

See Table 2 for childhood descriptive statistics among trajectory classes. Fifty-three percent of participants (*n* = 111) had complete data on the measurements used in the multinomial logistic regression. The regression model was initially estimated using an interaction term (i.e., caregiver education and race) based on previous findings within this sample (Bishop et al., 2007; Carr & Lord, 2012). However, the interaction was not statistically significant (*p* = .272). The full model was statistically significant [χ^2^(24)=83.78, *p* <.001]. Chi-square goodness-of-fit indicated that the model fit the data well [χ^2^(196)=173.04, *p* =.880]. Simple effects of multiple degrees of freedom tests revealed caregiver education (*p* =<.001), race (*p* =.04), ABC-I (*p* =.002), ABC-H (*p* =.03), and social supports (*p* =.01) all contributed significantly to the prediction of trajectory class membership. Table 3 further depicts the log-odds of belongingness to the medium and high negative impact classes relative to the low negative impact class across predictors. Nonparametric analyses revealed that racial minorities reported significantly lower ratings of objective and subjective burden in all CAIA domains compared to White participants.

**Table 2.** Predictor variables for trajectory classes

|  | Low Impact | Medium Impact | High Impact |  |
| --- | --- | --- | --- | --- |
| Child characteristics (*M, SD*) |  |  |  | *p*-value<sup>a</sup> |
| CSS SA | 5.32 (2.91) | 6.40 (2.70) | 6.36 (2.73) | .05 |
| CSS RRB | 5.77 (3.12) | 6.57 (2.81) | 7.62 (2.00) | .01 |
| ADI RRB | 3.92 (2.95) | 5.18 (2.80) | 6.50 (2.74) | <.01 |
| VABS DL | 62.09 (33.80) | 45.95 (28.56) | 39.74 (25.11) | <.01 |
| ABC Hyperactivity | 9.61 (9.38) | 14.31 (9.95) | 20.57 (11.09) | <.01 |
| ABC Irritability | 5.67 (8.00) | 10.58 (8.32) | 15.54 (11.14) | <.01 |
| VIQ | 65.43 (38.50) | 52.40 (37.81) | 46.89 (34.59) | .05 |
| Child (%) |  |  |  | *p*-value<sup>b</sup> |
| Sex (*Male*) | 80.88 | 75.51 | 88.37 | .21 |
| Race (*White*) | 60.29 | 82.65 | 90.70 | <.01 |
| ASD Diagnosis | 69.12 | 78.57 | 90.70 | .03 |
| Caregiver (%) |  |  |  |  |
| Marital Status | 75.86 | 79.57 | 81.40 | .75 |
| College Educated | 33.82 | 64.29 | 55.81 | <.01 |
| Social Support | 93.75 | 60.00 | 51.52 | <.01 |

*Note.* *p* value<sup>a</sup> = Analysis of Variance (ANOVA) with post-hoc comparisons; *p* value<sup>b</sup> = Chi-Square Test of Independence comparisons

**Table 3.** Multinomial logistic model for predicting negative impact trajectory class membership

| Trajectory Class | Predictors | $\beta$ | $p$-value | 95% Confidence Interval for<br>Exp $\beta$ | |
| --- | --- | --- | --- | --- | --- |
|  |  |  |  | Lower Bound | Upper Bound |
| **Medium Impact** | Caregiver Education | 3.20 | <.01 | 4.64 | 130.88 |
|  | Race | 1.40 | .15 | .59 | 27.82 |
|  | Marital Status | -1.04 | .31 | .05 | 2.61 |
|  | ABC Irritability | .31 | .01 | 1.09 | 1.71 |
|  | ABC Hyperactivity | -.17 | .02 | .73 | .97 |
|  | ADI-R RRB | .16 | .28 | .88 | 1.54 |
|  | ASD | -1.66 | .18 | .02 | 2.18 |
|  | ADOS-CSS SA | .17 | .28 | .87 | 1.63 |
|  | ADOS- CSS RRB | .06 | .69 | .78 | 1.46 |
|  | VIQ | -.01 | .50 | .95 | 1.02 |
|  | Social Support | 2.18 | .02 | 1.42 | 55.44 |
|  | VABS DL | -.01 | .84 | .95 | 1.04 |
| **High Impact** | Caregiver Education | 1.74 | .07 | .87 | 37.32 |
|  | Race | 2.90 | .02 | 1.59 | 206.33 |
|  | Marital Status | -.16 | .90 | .08 | 9.57 |
|  | ABC Irritability | .31 | .01 | 1.09 | 1.73 |
|  | ABC Hyperactivity | -.14 | .10 | .75 | 1.02 |
|  | ADI-R RRB | .38 | .03 | 1.04 | 2.06 |
|  | ASD | -1.03 | .51 | .02 | 7.85 |
|  | ADOS-CSS SA | .03 | .87 | .70 | 1.53 |
|  | ADOS- CSS RRB | .35 | .14 | .89 | 2.26 |
|  | VIQ | -.03 | .28 | .93 | 1.02 |
|  | Social Support | 2.80 | .01 | 2.17 | 125.27 |
|  | VABS DL | <-.01 | .94 | .95 | 1.05 |

### Trajectory Class and Number of Positive Outcomes

Due to attrition, functional outcome data were not available for the entire sample; thus, these analyses were conducted on a subset (*n =* 129). Ordinal regressions revealed that trajectory class was significantly associated with the number of positive outcomes adults achieved (*p* = .003). Higher odds of achieving more functional outcomes were associated with membership in the low impact class relative to the two other classes (β = -0.71, Odds Ratio [*OR*]= .49, *SE*= 0.24, *p* = .003).

## Discussion

The current study identified three distinct trajectory classes of parental perceived negative impact in a longitudinal cohort of participants with and without ASD. The largest class peaked in parental perceived negative impact when their son/daughter was an adolescent before decreasing into emerging adulthood. This finding is consistent with previous literature, which found that caregiver burden increases as children reach adolescence, possibly due to psychological, physiological, and social changes (Anderson, 2008; Carr & Lord, 2012; Corbett & Simon 2014; Gray, 2002; Rodriguez et al., 2019). In contrast, over half of the sample experienced low or high negative impact that steadily decreased over time. While overall findings indicated that parents reported less perceived negative impact as their children grew into emerging adulthood, their feelings of burden never completely subsided. Moreover, the high negative impact class retained scores that remained at least eight points higher than any other class across time These findings highlight the importance of understanding factors that contribute to parental burden as children with autism age into adults, particularly given that many parents remain primary caregivers for many, many years (Howlin & Magiati, 2017; Marsack & Hopp, 2019; Marsack-Topolewski et al., 2021). Moreover, increased caregiver burden is related to lower levels of quality of life for aging parents of adult children with ASD (Marsack-Topolewski & Church, 2019).

Core autism features and child cognitive abilities did not affect parents’ perceptions of burden into emerging adulthood, in contrast to several other child and family factors such as hyperactivity, irritability, social support, race, and caregiver education. These findings replicate our previous analyses of these data during childhood and adolescence (Bishop et al., 2007; Carr & Lord, 2012). They are also similar to the literature finding a clear relationship between externalizing symptoms in children and increased parental stress (Barroso et al., 2018; Lecavalier et al., 2006; Sui et al., 2019; Yorke et al., 2018; Zaidman-Zait et al., 2014). Our sample found that caregivers who had children with increased hyperactivity and irritability symptoms were likely to be members of the highest perceived negative impact class. Therefore, as children are diagnosed with ASD, hyperactivity and irritability should be continually assessed and monitored (Ip et al., 2019). As others have noted (Lamy & Erickson, 2018), treatments are available for co-occurring hyperactivity (Antshel & Russo, 2019) and irritability (Bartram et al., 2019; Behmanesh et al., 2019). Other proactive strategies like coping and mindfulness may help caregivers manage difficult stressors to improve long-term functional outcomes in adulthood (Cachia et al., 2016).

While it is important to understand the predictive variables, each trajectory class’s phenotypic makeup has potential implications. First, participants in the high negative impact class exhibited lower daily living adaptive skills relative to the other two classes. Daily living skills are somewhat responsive to treatment, suggesting that increased support for families in this area might not only improve adaptive skills but result in less negative impact on caregivers (Bal et al., 2015; Duncan & Bishop, 2015; Drahota, 2011). Without a larger sample size, it is not possible to statistically evaluate mediators, but future studies could address the degree to which the relationship of perceived negative impact was mediated by daily living skills. Second, individuals in the low negative impact class attained more positive functional outcomes as adults relative to the other classes, controlling for IQ. It may be that caregivers who manage burden well may feel more able to support positive outcomes.

Lastly, the current study found that participants belonging to the high negative impact class exhibited more clinically observed and caregiver reported RRBs. Children who spend large amounts of time engaging in RRBs may be more difficult to interact with and less socially connected with others around them (Harrop et al., 2014; Jones et al., 2017; Loftin et al., 2008). As with hyperactivity, irritability, and adaptive skills, RRBs affect not only the child but the families who care for them.

Parental characteristics, including higher education and being White, were predictive of membership in the high negative impact class. This contrasts with the parental stress literature, where racial minority families of autistic children have typically reported higher stress levels than White families (DeLambo et al., 2011; Kim et al., 2020; Williams et al., 2019). Potential reasons for lower ratings of parental perceived negative impact in minority families in this study include stronger coping skills (including religion), attribution of feelings of burden to other sources (not related to the child with a disability), strong family resilience, and acculturation (Bishop et al., 2007; Carr & Lord, 2012; Kim et al., 2020; Williams et al., 2019). At each point during our assessments, a third of our interviewers were racial minorities, but we did not make an effort to match examiner and participant race. It is possible that racial minority parents were less comfortable reporting concerns to a White researcher. Another possible explanation is that racial minorities report less impact related to financial, leisure, work, social, and other objective and subjective burdens that comprise negative impact than typical measures of stress, where the focus is often on subjective feelings about children’s behaviors.

Caregivers who felt socially supported more often belonged to the low negative impact class, a finding consistent with past research (Bishop et al., 2007; Carr & Lord, 2012; Goedeke et al., 2019; Marsack & Samuel, 2017; Zaidman-Zait et al., 2017). This is another factor that may often get lost during typical diagnostic assessments or medical management that often focus on autism or psychiatric symptoms. It is important that clinicians inquire about caregiver supports (i.e., partner, family, friend, professional) and assist families with accessing resources to reduce parental burden and stress and increase the likelihood of positive adult outcomes for those with ASD.

### Strengths and Limitations

A significant strength of this study is its longitudinal design, which provides insight into how parental perceived negative impact changes over time. Most literature, including this study, focuses on the mother’s perceptions of negative impact, which is both a strength and limitation of this study. Note that this study did not focus on the positive experiences of raising a child with autism, which future studies could explore in greater depth. Although this study had attrition, we were able to retain African-American families as 20 percent of the sample, which is actually higher than the proportion of American-Americans in these geographic locations. Future studies might explore potential mediators or moderators of parental perceived negative impact (i.e., coping skills, discrimination experience, parental self-efficacy, attributions, acculturation, family resilience, etc.) and define caregiver outcomes (i.e., well-being, mental health issues, happiness, quality of life) to further understand their relationship in perceived negative impact.

Despite these limitations, the present study contributes several new and important findings. First, we found different trajectories of perceived negative impact over time, all of which never completely subsided into early adulthood. These findings highlight the need to provide resources to support and alleviate the burden in aging parents of adults with ASD (Marsack-Topolewski & Church, 2019). Family factors like minority status and lower education level were related to those families with lower levels of perceived negative impact that steadily decreased with time. Those belonging to the largest group, the medium negative impact class, rating of burden increased as their children entered adolescence. Caregivers in the high impact class steadily decreased with time, but overall remained very high compared to the other classes. Second, treatable child characteristics like hyperactivity and irritability predicted class membership, rather than less malleable characteristics (e.g., intelligence and autism severity).

We also found caregivers who felt socially supported belonged to lower impact classes. This finding is not surprising but yet clinically relevant. Lastly, the study showed a relationship between the amount of caregiver burden and the number of functional outcomes attained by adults with ASD (i.e., increased parental burden was associated with adults meeting less functional outcomes). Unfortunately, the directionality of the relationship was not examined, but those who achieved more functional outcomes as adults demonstrated higher daily living skills. Daily livings skills have been well-documented as a deficit in those with ASD. Given the lack of research on adults with autism and their caregivers, future studies should focus on the needs, quality of life, and well-being of aging caregivers supporting their adult child with ASD.

## Data Availability

All data produced in the present study are available upon reasonable request to the authors.

## References

Aman, M., Singh, N., Stewart, A., & Field, C. (1985). The Aberrant Behavior Checklist: A behavior rating scale for the assessment of treatment effects. American Journal of Mental Deficiency, 89, 485–491.

Anderson, L. (2008). Predictors of parenting stress in a diverse sample of parents of early adolescents in high-risk communities. Nursing Research, 57(5), 340–350. https://doi.org/10.1097/01.NNR.0000313502.92227.87

Angold, A., Messer, S., Stangl, D., Farmer, E., Costello, E., & Burns, B. (1998). Perceived parental burden and service use for child and adolescent psychiatric disorders. American Journal of Public Health, 88(1), 75–80. https://ajph.aphapublications.org/doi/pdfplus/10.2105/AJPH.88.1.75

Antshel, K., & Russo, N. (2019). Autism spectrum disorders and ADHD: Overlapping phenomenology, diagnostic issues, and treatment considerations. Current Psychiatry Report, 21(5), 1–11. https://doi.org/10.1007/s11920-019-1020-5

Bal, V., Kim, S., Cheong, D., & Lord, C. (2015). Daily living skills in individuals with autism spectrum disorder from 2 to 21 years of age. Autism, 19(7), 774–784. https://doi.org/10.1177/1362361315575840

Barroso, N.E., Mendez, L., Graziano, P.A., & Bagner, D. (2018). Parenting stress through the lens of different clinical groups: A systematic review & meta-analysis. Journal Abnormal Child Psychology, 46(3), 449–461. https://doi.org/10.1007/s10802-017-0313-6

Bartram, L., Lozano, J., & Coury, D. (2019). Aripiprazole for treating irritability associated with autism spectrum disorders. Expert Opinion on Pharmacotherapy, 20(12), 1421–1427. https://doi.org/10.1080/14656566.2019.1626825

Behmanesh, H., Moghaddam, H., Mohammadi, M., & Akhondzadeh, S. (2019). Risperidone combination therapy with propentofylline for treatment of irritability in autism spectrum disorders: A randomized, double-blind, placebo-controlled clinical trial. Clinical Neuropharmacology, 42(6), 189–196. https://doi.org/10.1097/WNF.0000000000000368

Bishop, S., Richler, J., Cain. C., & Lord, C. (2007). Predictors of perceived negative impact in mothers of children with autism spectrum disorder. American Journal of Mental Retardation, 112(6), 450–461. https://doi.org/10.1352/08958017(2007)112[450:POPNII]2.0.CO;2

Bonis, S. (2016). Stress and parents of children with autism: A review of literature. Issues in Mental Health Nursing, 37(3), 153–163. https://doi.org/10.3109/01612840.2015.1116030

Carr, T. & Lord, C. (2013). Longitudinal study of perceived negative impact in African American and Caucasian mothers of children with autism spectrum disorder. Autism, 17(4), 405–417.

Cachia, R., Anderson, A., & Moore, D. (2015). Mindfulness, stress, and well-being in parents of children with autism spectrum disorder: a systematic review. Journal of Child and Family Studies, 25(1), 1–14. doi:10.1007/s10826-015-0193-8.

Corbett, B., & Simon, D. (2014). Adolescence, stress and cortisol in autism spectrum disorders. O.A. Autism, 1(1), 2–11. https://www.ncbi.nlm.nih.gov/pmc/articles/PMC3961758/

Costello, E., Angold, A., Burns, B., Stangl, D., Tweed, D., Erkanli, E., & Worthman, C. (1996) The Great Smoky Mountains study of youth: goals, designs, methods, and the prevalence of DSM-III-R disorders. Archives General Psychiatry, 53(12), 1129–1136. https://doi.org/110.1001/archpsyc.1996.01830120067012

Crum, K.I., & Moreland, A.D. (2017). Parental Stress and Children’s Social and Behavioral Outcomes: The Role of Abuse Potential over Time. Journal Child Family Studies, 26, 3067–3078. https://doi.org/10.1007/s10826-017-082

DeLambo, D., Chung, W., & Huang, W. (2011). Stress and age: A Comparison of Asian American and non-Asian American parents of children with developmental disabilities. Journal of Developmental & Physical Disabilities, 23(2), 129–141. https://doi.org/10.1007/s10882-010-9211-3

DesChamps, T., Ibañez, L., Edmunds, S., Dick, C., & Stone, W. (2020). Parenting stress in caregivers of young children with ASD concerns prior to a formal diagnosis. Autism Research, 13(1), 82–92. https://doi.org/10.1002/aur.2213

Drahota, A., Wood, J., Sze, K., & Van Dyke, M. (2011). Effects of cognitive behavioral therapy on daily living skills in children with high-functioning autism and concurrent anxiety disorders. Journal of Autism and Developmental Disorders, 41(3), 257–265. https://doi.org/10.1007/s10803-010-1037-4

Duncan, A., & Bishop, S. (2013). Understanding the gap between cognitive abilities and daily living skills in adolescents with autism spectrum disorders with average intelligence. Autism, 19(1), 64–72. https://doi.org/10.1177/1362361313510068

Elliott, C. D., Murray, G. J., & Pearson, L. S. (1990). Differential ability scales. San Antonio, Texas.

Estes, A., Olson, E., Sullivan, K., Greenson, J., Winter, J., Dawson, G., & Munson, J. (2013). Parenting-related stress and psychological distress in mothers of toddlers with autism spectrum disorders. Brain & Development, 35(2), 133–138. https://doi.org/10.1016/j.braindev.2012.10.004

Goedeke, S., Shepherd, D., Landon, J., & Taylor, S. (2019). How perceived support relates to child autism symptoms and care-related stress in parents caring for a child with autism. Research in Autism Spectrum Disorders, 60, 36–47. https://doi.org/10.1016/j.rasd.2019.01.005

Gray, D. (2002). Ten years on: a longitudinal study of families of children with autism. Journal of Intellectual and Developmental Disability, 27(3), 215–222. https://doi.org/10.1080/1366825021000008639

Harrop, C., Gulsrud, A., Shih, W., Hovsepyan, L., & Kasari, C. (2017). The impact of caregiver mediated JASPER on child restricted and repetitive behaviors and caregiver responses. Autism Research, 10(5), 983–992. https://doi.org/10.1002/aur.1732

Hayes, S. A., & Watson, S. L. (2013). The impact of parenting stress: A meta-analysis of studies comparing the experience of parenting stress in parents of children with and without autism spectrum disorder. Journal of Autism and Developmental Disorders, 43(3), 629–642. https://doi.org/10.1007/s10803-012-1604-y

Howlin, P., Moss, P., Savage, S., & Rutter, M. (2013). Social outcomes in mid-to later adulthood among individuals diagnosed with autism and average nonverbal IQ as children. Journal American Academy Child and Adolescent Psychiatry, 52(6), 572–8. doi: 10.1016/j.jaac.2013.02.017. Epub 2013 Apr 24. PMID: 23702446.

Howlin, P., & Magiati, I. (2017). Autism spectrum disorder: Outcomes in adulthood. Current Opinion in Psychiatry, 30(2), 69–76. https://doi.org/10.1097/YCO.0000000000000308

Ip, A., Zwaigenbaum, L., & Brian, J. (2019). Post-diagnostic management and follow-up care for autism spectrum disorder. Pediatrics & Child Health, 24(7), 461–468. https://doi.org/10.1093/pch/pxz121

Jones, B., Nagin, D., & Roeder, K. (2001). A SAS procedure based on mixture models for estimating developmental trajectories. Sociological Methods & Research, 29(3), 374–393. https://doi.org/10.1177/0049124101029003005

Jones, B., & Nagin, D. (2007). Advances in group-based trajectory modeling and an SAS procedure for estimating them. Sociological Methods & Research, 35(4), 542–571. https://doi.org/10.1177/0049124106292364

Jones, B., & Nagin, D. (2013). A note on a Stata plugin for estimating group-based trajectory models. Sociological Methods & Research, 42(4), 608–613. https://doi.org/10.1177/0049124113503141

Jones, J.H., Call, T.A., Wolford, S.N., & McWey, M. (2021). Parental Stress and Child Outcomes: The Mediating Role of Family Conflict. Journal of Child Family Studies, 30, 746–756. https://doi.org/10.1007/s10826-021-01904-8

Jones, R. M., Pickles, A., & Lord, C. (2017). Evaluating the quality of peer interactions in children and adolescents with autism with the Penn Interactive Peer Play Scale (PIPPS). Molecular Autism, 8(28), 1–9. https://doi.org/10.1186/s13229-017-0144-x

Kayfitz, A. D., Gragg, M. N., & Orr, R. R. (2010). Positive experiences of mothers and fathers of children with autism. Journal of Applied Research in Intellectual Disabilities, 23,337–343. https://doi.org/10.1111/j.1468-3148.2009.00539.x

Kim, I., Dababnah, S., & Lee, J. (2020). The influence of race and ethnicity on the relationship between family resilience and parenting stress in caregivers of children with autism. Journal of Autism and Developmental Disorders, 50(2), 650–658. https://doi.org/10.1007/s10803-019-04269-6.

King, G., Zwaigenbaum, L., Bates, A., Baxter, D., Rosenbaum, P. (2012). Parent views of the positive contributions of elementary and high school-aged children with autism spectrum disorders and Down syndrome. Child: Care, Health, and Development, 38, 817–828. doi:10.1111/j.1365-2214.2011.01312.x

Lamy, M., & Erickson, C. (2018). Pharmacological management of behavioral disturbances in children and adolescents with autism spectrum disorders. Current Problems Pediatric Adolescent Health Care, 48(10), 250–264. https://doi.org/10.1016/j.cppeds.2018.08.015

Lecavalier, L., Leone, S., & Wiltz, J. (2006). The impact of behaviour problems on caregiver stress in young people with autism spectrum disorders. Journal of Intellectual Disability Research, 50(3), 172–183. https://doi.org/10.1111/j.1365-2788.2005.00732.x

Loftin, R. L., Odom, S. L., & Lantz, J. F. (2008). Social interaction and repetitive motor behaviors. Journal of Autism and Developmental Disorders, 38(6), 1124–1135. https://doi.org/10.1007/s10803-007-0499-5

Lord, C., Rutter, M., & Le Couter, A. (1994). Autism diagnostic interview–revised: A revised version of a diagnostic interview for caregivers of individuals with possible pervasive developmental disorders. Journal of Autism and Developmental Disorders, 24(5), 659–685.

Lord, C., Rutter, M., DiLavore, P.C., Risi, S., Gotham, K., & Bishop, S.L. (2012). Autism diagnostic observation schedule, 2nd edition (ADOS-2), Los Angeles, CA: Western Psychological Corporation.

Lounds Taylor, J. (2017). When is a good outcome actually good? Autism, 21(8), 918–919. https://doi.org/10.1177/1362361317728821

Marsack, C. N., & Samuel, P. S. (2017). Mediating Effects of Social Support on Quality of Life for Parents of Adults with Autism. Journal of autism and developmental disorders, 47(8), 2378–2389. https://doi.org/10.1007/s10803-017-3157-6

Marsack, C. N., & Hopp, F. P. (2019). Informal Support, Health, and Burden Among Parents of Adult Children With Autism. The Gerontologist, 59(6), 1112–1121. https://doi.org/10.1093/geront/gny082

Marsack-Topolewski, C. & Church, H. (2019). Impact of caregiver burden on quality of life for parents of adult children with Autism Spectrum Disorder. American Journal on Intellectual and Developmental Disabilities, 124(2):145–156. doi: 10.1352/1944-7558-124.2.145. PMID: 30835531.

Marsack-Topolewski, C., Samuel, P., & Tarraf, W. (2021) Empirical evaluation of the association between daily living skills of adults with autism and parental caregiver burden. Plos One, 16(1), https://doi.org/10.1371/journal.pone.0244844

McCauley, J., Pickles, A., Huerta, M., & Lord, C. (2020). Defining positive outcomes in more and less cognitively able autistic adults. Autism Research, 13(9), 1548–1560. https://doi.org/10.1002/aur.2359

McStay, R. L., Trembath, D., & Dissanayake, C. (2014). Stress and family quality of life in parents of children with autism spectrum disorder: Parent gender and the double ABCX model. Journal of Autism and Developmental Disorders, 44(12), 3101–3118. https://doi.org/10.1007/s10803-014-2178-7

Messer, S., Angold, A., Costello, E., & Burns, B. (1996). The Child and Adolescent Burden Assessment (CABA): Measuring the family impact of emotional and behavioral problems. International Journal of Methods in Psychiatric Research, 6(4), 261–284. https://doi.org/10.1002/(SICI)1234-988X(199612)6:4<261::AID-MPR169>3.3.CO;2-3

Mullen, E. (1995). The Mullen Scales of Early Learning. Circle Pines, MN: American Guidance Service.

Nagin, D. (2005). Group-based modeling of development Harvard University Press. Cambridge, MA.

Pastor-Cerezuela, G., Fernández-Andrés, M. I., Tárraga-Minguez, R., & Navarro-Peña, J. M. (2015). Parental stress and ASD: Relationship with autism symptom severity, IQ, and resilience. Focus on Autism and Other Developmental Disabilities, 31(4), 300–311. https://doi.org/10.1177/1088357615583471

Picardi, A., Gigantesco, A., Tarolla, E., Stoppioni, V., Cerbo, R., Cremonte, M., Alessandri, G., Lega, I., & Nardocci, F. (2018). Parental burden and its correlates in families of children with autism spectrum disorder: A multicentre study with two comparison groups. Clinical Practice and Epidemiology in Mental Health: C.P. & EMH, 14, 143–176. https://doi.org/10.2174/1745017901814010143

Pickles, A., McCauley, J. B., Pepa, L. A., Huerta, M., & Lord, C. (2020). The adult outcome of children referred for autism: Typology and prediction from childhood. Journal of Child Psychology and Psychiatry, 61(7), 760–767. https://doi.org/10.1111/jcpp.13180

Raina, P., O’Donnell, M., Schwellnus, H. et al. (2004). Caregiving process and caregiver burden: Conceptual models to guide research and practice. BMC Pediatrics, 4, 1 https://doi.org/10.1186/1471-2431-4-1

Rodriguez, G., Hartley, S., & Bolt, D. (2019). Transactional relations between parenting stress and child autism symptoms and behavior problems. Journal of Autism and Developmental Disorders, 49(5), 1887–1898. https://doi.org/10.1007/s10803-018-3845-x

Shine, R., & Perry, A. (2010). Brief Report: The Relationship Between Parental Stress and Intervention Outcome of Children with Autism. Journal of Developmental Disabilities, 16, 64–67

Siu, Q., Yi, H., Chan, R., Chio, H., Chan, F., & Mak, W. (2019). The role of child problem behaviors in autism spectrum symptoms and parenting stress: A primary school-based study. Journal of Autism and Developmental Disorders, 49(3), 857–870. https://doi.org/10.1007/s10803-018-3791-7

Sparrow S., Cicchetti, V., & Balla A. (2005). Vineland adaptive behavior scales. 2nd. Circle Pines, MN: American Guidance Service

Stringer, D., Kent, R., Briskman, J., Lukito, S., Charman, T., Baird, G., Lord, C., Pickles, A., & Simonoff, E. (2020). Trajectories of emotional and behavioral problems from childhood to early adult life. Autism, 24(4), 1011–1024. https://doi.org/10.1177/1362361320908972

Taylor, J., Henninger, N., & Mailick, M. (2015). Longitudinal patterns of employment and postsecondary educational activities for adults with ASD and normal-range I.Q. Autism, 19(7), 785–793. https://doi.org/10.1177/1362361315585643

Tsermentseli, S., & Kouklari, E. (2019). Impact of child factors on parenting stress of mothers of children with autism spectrum disorder and intellectual disability: a U.K. school-based study. Early Child Development and Care, 1–12. https://doi.org/10.1080/03004430.2019.1658090

Trute, B., & Hiebert-Murphy, D. (2005). Predicting family adjustment and parenting stress in childhood disability services using brief assessment tools. Journal of Intellectual and Developmental Disability, 30, 217–225. https://doi.org/10.1080/13668250500349441

U.S. Census Bureau (2020). Racial and Ethnic Diversity in the United States: 2010 Census and 2020 Census. Retrieved from https://www.census.gov/library/visualizations/interactive/racial-and-ethnic-diversity-in-the-united-states-2010-and-2020-census.html.

Wechsler, D. (1999). Wechsler Abbreviated Scale of Intelligence Psychological Corporation, San Antonio, TX: Psychological Corporation.

Wechsler, D. (2003). Wechsler Intelligence Scale for Children (4th ed.). San Antonio, TX: Harcourt Assessment.

Williams, T., Hartmann, K., Paulson, J., Raffaele, C., & Urbano, M. (2019). Life after an autism spectrum disorder diagnosis: A comparison of stress and coping profiles of African American and Euro-American caregivers. Journal Autism Developmental Disorder, 49(3), 1024–1034. https://doi.org/10.1007/s10803-018-3802-8

Yorke, I., Pippa, W., Weston, A., Rafla, M., Charman, T., & Simonoff, E. (2018). The association between emotional and behavioral problems in children with autism spectrum disorder and psychological distress in their parents: a systematic review and meta-analysis. Journal of Autism and Developmental Disorders, 48(10), 3393–3415. https://doi.org/10.1007/s10803-018-3605-y

Zaidman-Zait, A., Mirenda, P., Duku, E., Szatmari, P., Georgiades, S., & Volden, J., Zwaigenbaum, L., Vaillancourt, T., Bryson, S., Smith, I., Fombonne, E., Roberts, W., Waddell, C., Thompson, A., & & Pathways in ASD Study Team. (2014). Examination of bidirectional relationships between parent stress and two types of problem behavior in children with autism spectrum disorder. Journal of Autism and Developmental Disorders, 44(8), 1908–1917. https://doi.org/10.1007/s10803-014-2064-3

Zaidman-Zait, A., Mirenda, P., Duku, E., Vaillancourt, T., Smith, I. M., Szatmari, P., Bryson, S., Fombonne, E., Volden, J., Waddell, C., Zwaigenbaum, L., Georgiades, S., Bennett, T., Elsabaggh, M., & Thompson, A. (2017). Impact of personal and social resources on parenting stress in mothers of children with autism spectrum disorder. Autism, 21(2), 155–166. https://doi.org/10.1177/1362361316633033

